# Multi-Layered Dysregulation of NRCAM in Gliomas: Insights from TCGA Copy Number and Epigenetic Analyses

**DOI:** 10.1101/2025.08.19.25333982

**Authors:** Steven Lehrer, Peter H. Rheinstein

## Abstract

**Background:** NRCAM, a neuronal cell adhesion molecule, has been implicated in glioma biology through splicing alterations reported by prior studies. However, the relative contributions of genomic and epigenetic mechanisms to NRCAM dysregulation in gliomas remain unclear.

**Methods:** We analyzed glioma datasets from The Cancer Genome Atlas (TCGA) using UCSC Xena and cBioPortal. Copy number variation (CNV), DNA methylation (Illumina 450K arrays), and mutation profiles were assessed for NRCAM. Kaplan Meier survival analyses were performed with Xena, stratifying patients by copy number status and methylation state. Correlations between mutation burden and fraction genome altered were evaluated using Spearman and Pearson methods.

**Results:** Somatic mutations in NRCAM were rare across TCGA gliomas. In contrast, CNV and methylation changes were frequent and clinically relevant. Copy number gains at the NRCAM locus were associated with significantly shorter overall survival, while higher methylation of NRCAM correlated with improved survival outcomes. NRCAM mutation count did not show a linear correlation with fraction genome altered (FGA), suggesting these alterations are largely independent of overall genomic instability. The findings highlight copy number imbalance and epigenetic regulation as predominant mechanisms of NRCAM dysregulation.

**Conclusion:** NRCAM is recurrently dysregulated in gliomas through copy number alterations and DNA methylation, both of which stratify patient survival. Together with previously reported splicing deregulation, these data suggest that NRCAM functions as a multilayered regulator of glioma progression. NRCAM methylation may represent a prognostic biomarker, while therapeutic modulation of NRCAM warrants further investigation.

## Introduction

Glioblastoma and other high-grade gliomas remain among the most aggressive cancers, with limited therapeutic options and poor survival outcomes. Despite advances in molecular profiling, current biomarkers such as IDH mutation status and MGMT promoter methylation explain only part of the observed heterogeneity in patient prognosis. Large-scale initiatives such as The Cancer Genome Atlas (TCGA) have provided comprehensive catalogs of somatic mutations, copy number alterations, and DNA methylation patterns, enabling the identification of new molecular features that may contribute to disease biology and outcome.

One emerging area of interest in glioma biology is the role of alternative splicing [1, 2]. Microexons—very short exons of 3–27 nucleotides—are particularly enriched in neuronal tissues, where they regulate protein–protein interactions critical for synaptic development and neuronal signaling. Dysregulation of microexons has been implicated in neurodevelopmental disorders, and recent work suggests they may also contribute to tumorigenesis. For example, neuronal cell adhesion molecule (NRCAM), a member of the L1 family of immunoglobulin-like cell adhesion molecules, undergoes microexon regulation, and altered isoform has been linked to invasive and proliferative phenotypes in glioma models [3, 4].

Although these findings underscore the biological importance of NRCAM splicing, the contribution of genomic and epigenomic alterations at the NRCAM locus in human gliomas remains poorly defined. Unlike point mutations in oncogenes and tumor suppressors, which are well characterized in glioma, structural and regulatory changes in genes such as NRCAM have received comparatively little attention. It is not yet clear whether copy number variation, DNA methylation, or other locus-specific changes are associated with clinical outcomes, nor how these alterations might intersect with broader glioma biology.

The objective of this study is to address this knowledge gap by performing a systematic evaluation of NRCAM alterations in TCGA glioma cohorts. By integrating analyses of copy number, mutation, and methylation data, we aimed to determine whether this locus exhibits reproducible patterns of dysregulation that stratify patients by prognosis. Our goal is to establish whether NRCAM genomic and epigenomic features may serve as clinically relevant biomarkers in glioma and related tumors, thereby extending the scope of glioma molecular profiling beyond established drivers.

## Methods

Data were obtained from TCGA GDC Lower Grade Glioma (LGG) cohort [5]. Publicly available datasets were accessed through UCSC Xena (https://xena.ucsc.edu/) [6] and cBioPortal for Cancer Genomics (https://www.cbioportal.org/) [7]. These platforms provide TCGA datasets for copy number variation (CNV), DNA methylation, somatic mutations, and clinical outcomes.

Copy number data were downloaded from UCSC Xena, which hosts segmented CNV calls derived from multiple algorithms including GISTIC2 and ASCAT. GISTIC2 was used to identify recurrently altered regions, while ASCAT was used to infer allele-specific copy numbers and overall FGA for each tumor sample. GISTIC thresholds refer to the cutoff values used in the GISTIC algorithm (Genomic Identification of Significant Targets in Cancer) to classify the degree of copy number alterations (CNAs) across the genome [8]. ASCAT stands for Allele-Specific Copy number Analysis of Tumors, a computational algorithm developed to infer Allele-specific copy number changes (i.e., distinguishing between maternal and paternal alleles), tumor purity (the proportion of tumor cells in the sample versus normal cells), and tumor ploidy (average number of chromosome sets per tumor cell). ASCAT takes SNP array or sequencing intensity data and adjusts for normal cell admixture and aneuploidy, allowing a more accurate determination of copy number variations (CNVs) in cancer genomes. ASCAT is widely used in TCGA and other large cancer genomics projects to generate fraction genome altered (FGA) and copy number segment profiles. CNV status at the NRCAM locus was extracted for each patient.

DNA methylation data were obtained from the Illumina HumanMethylation450 BeadChip arrays and displayed on UCSC Xena. β-values for probes mapping to the NRCAM gene locus were extracted. Both individual probes and aggregate measures were examined to assess methylation status. The median was calculated across all samples in the cohort to define the high and low methylation groups.

Somatic mutation data were obtained from Xena and cBioPortal. Mutation frequencies and types within the NRCAM gene were catalogued.

Clinical outcome data, including overall survival (OS) and progression-free survival (PFS), were obtained from UCSC Xena. Kaplan–Meier survival curves were generated directly within the UCSC Xena browser by stratifying patients according to NRCAM CNV status or methylation groupings (median split of β-values). Survival differences were evaluated using log-rank tests provided by the Xena interface.

## Results

Figure 1 shows characteristics of 495 subjects in the TCGA GDC Lower Grade Glioma (LGG) cohort: Age distribution versus number of cases, Sex distribution, Racial distribution, Overall survival (months).

**Figure 1.**
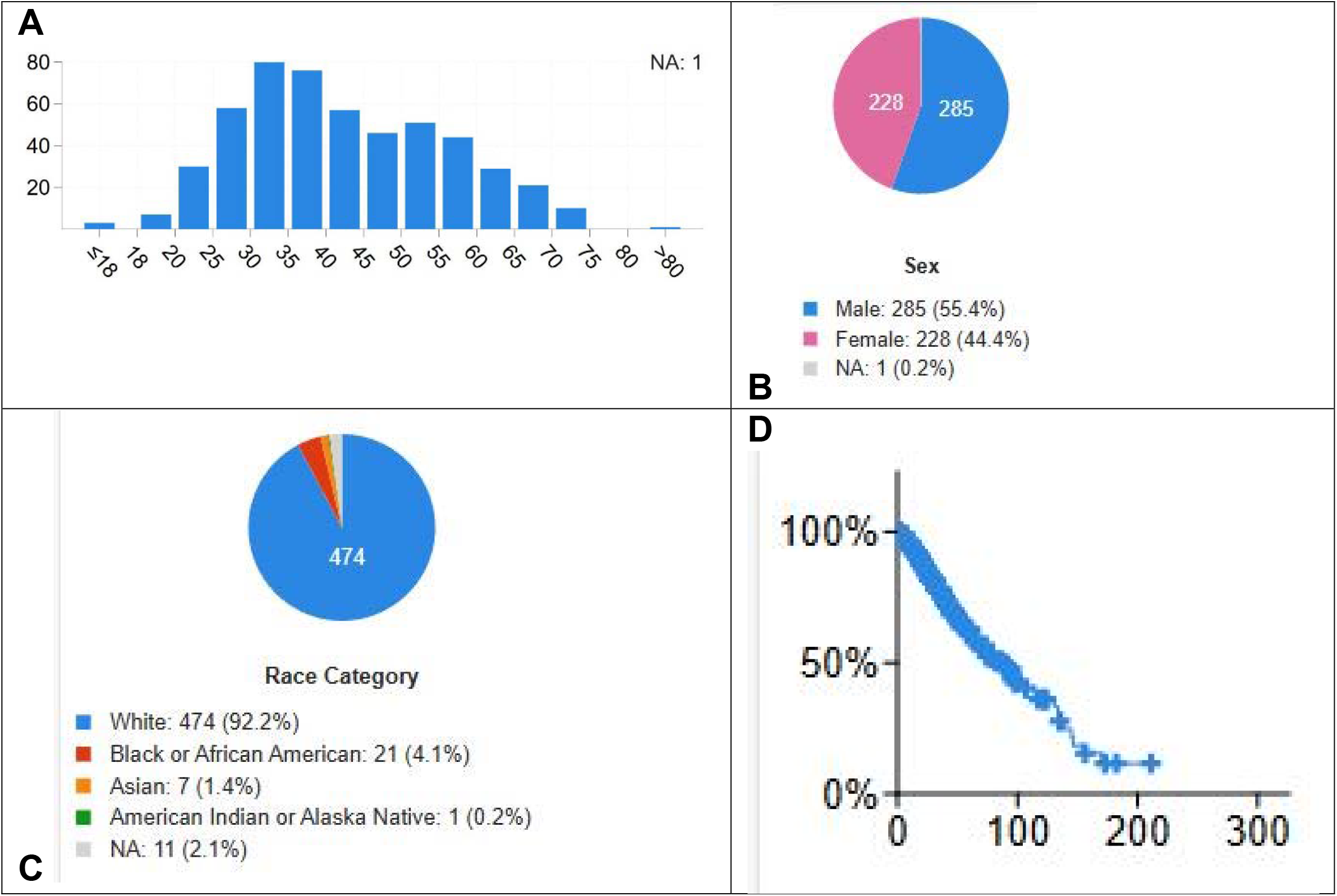
TCGA GDC Lower Grade Glioma (LGG) cohort. 1A Age distribution versus number of cases. 1B Sex distribution. 1C Racial distribution. 1D. Overall survival (months).

Figure 2 shows NRCAM expression in the GDC TCGA Lower Grade Glioma (LGG) dataset visualized using UCSC Xena Browser. 4 NRCAM missense mutations and 4 amplifications were present in 495 subjects. NRCAM lower copy number is associated with the 1p 19q co-deletion, absence of TP53 and ATRX mutations in lower grade tumors, and better survival (Figure 3). Primary diagnosis terms are from the old classification system [9].

**Figure 2.**
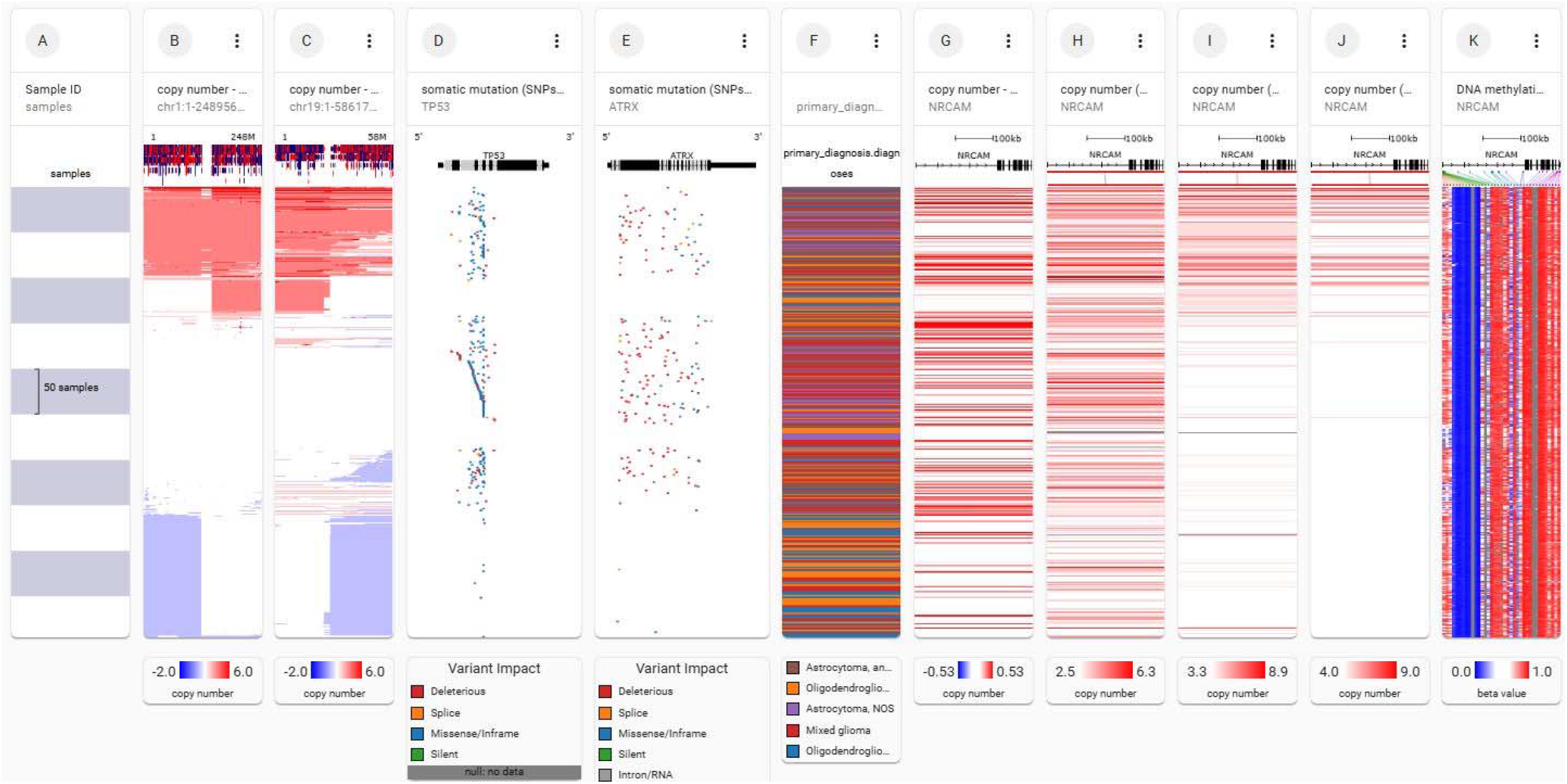
NRCAM expression in the GDC TCGA Lower Grade Glioma (LGG) dataset visualized using UCSC Xena Browser. Each row contains data from a single sample. Row order is determined by sorting the rows by their column values. Red indicates a copy number gain or amplification. The more intense the red, the higher the copy number. Blue indicates a copy number loss or deletion. The more intense the blue, the greater the loss. White represents a normal copy number (diploid state). This composite visualization displays data from 495 subjects. (A) Sample index. (B) Allele specific copy number chr 1 (ASCAT 3) (C) Allele specific copy number chr 19 (ASCAT 3). B and C show the 1p 19q codeletion (vertical blue bars). (D) Somatic mutations TP53. (E) Somatic mutations ATRX. (F) Primary Diagnosis. (G) NRCAM copy number masked copy number segment (DNA copy). (H) NRCAM gene level copy number absolute (I) NRCAM gene level copy number ASCAT 2 (J)NRCAM gene level copy number ASCAT 3 (J) NRCAM DNA methylation Illumina methylation 450.

**Figure 3.**
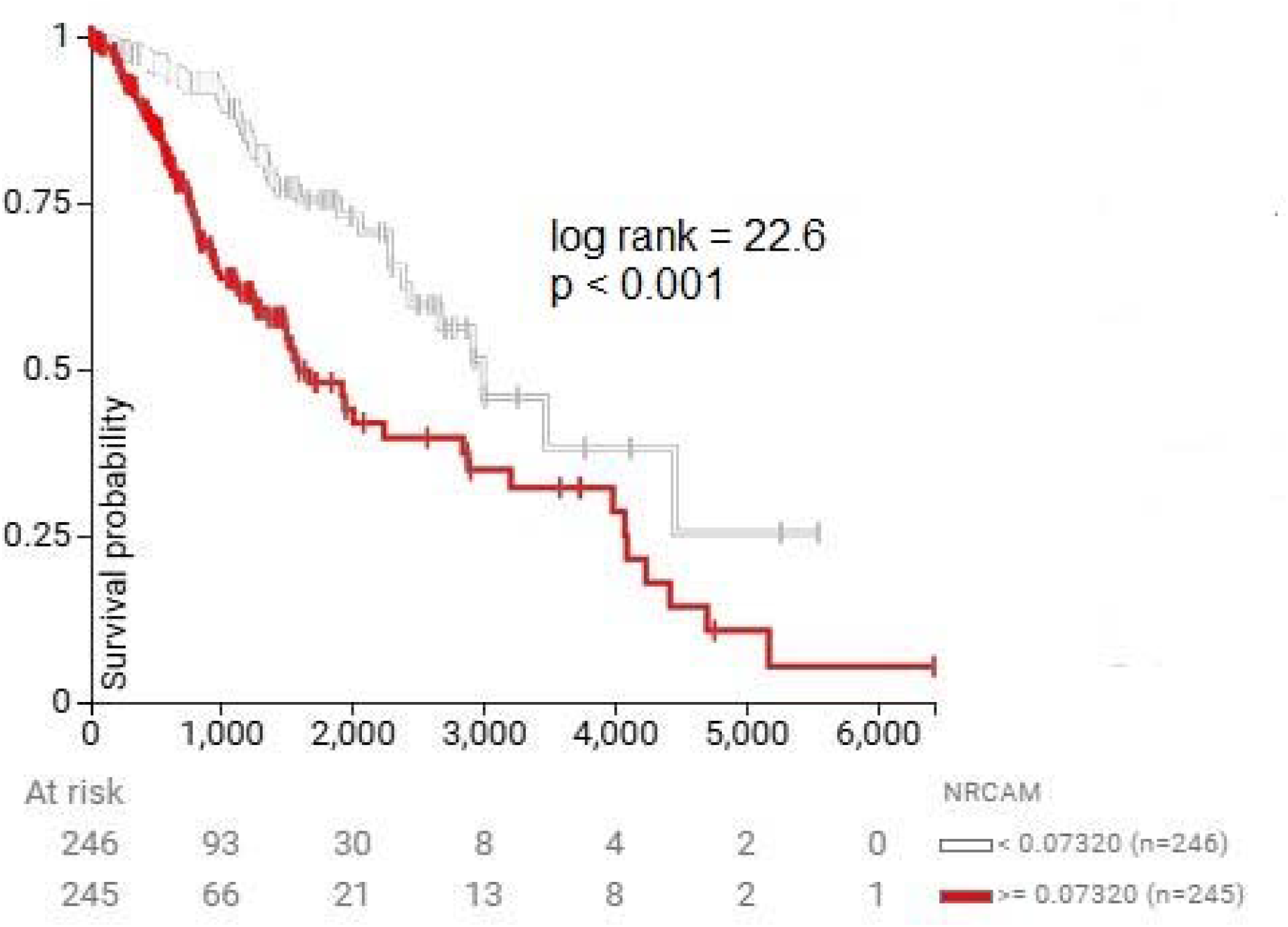
Survival probability 491 subjects stratified by NRCAM copy number. Subjects with lower copy number (upper curve) had better survival.

Somatic mutation data obtained through Xena and cBioPortal demonstrated that NRCAM mutations were rare in gliomas. Four alterations were missense mutations of uncertain functional significance; four others were amplifications. Mutations rarely co-occurred with high-level copy number changes at the same locus.

Kaplan–Meier survival analyses were conducted directly in UCSC Xena. Patients were stratified according to NRCAM copy number status and methylation levels. Survival curves indicated that NRCAM copy number gains were generally associated with shorter overall survival (Figure 3).

DNA methylation data from Illumina HumanMethylation450 arrays showed variable methylation across multiple probes mapped to the NRCAM gene. Stratification of cases by median β-values revealed two distinct groups: low-methylation and high-methylation tumors. Methylation states were not evenly distributed across glioma subtypes; higher methylation was more frequently observed in lower-grade gliomas with increased survival (Figure 4).

**Figure 4.**
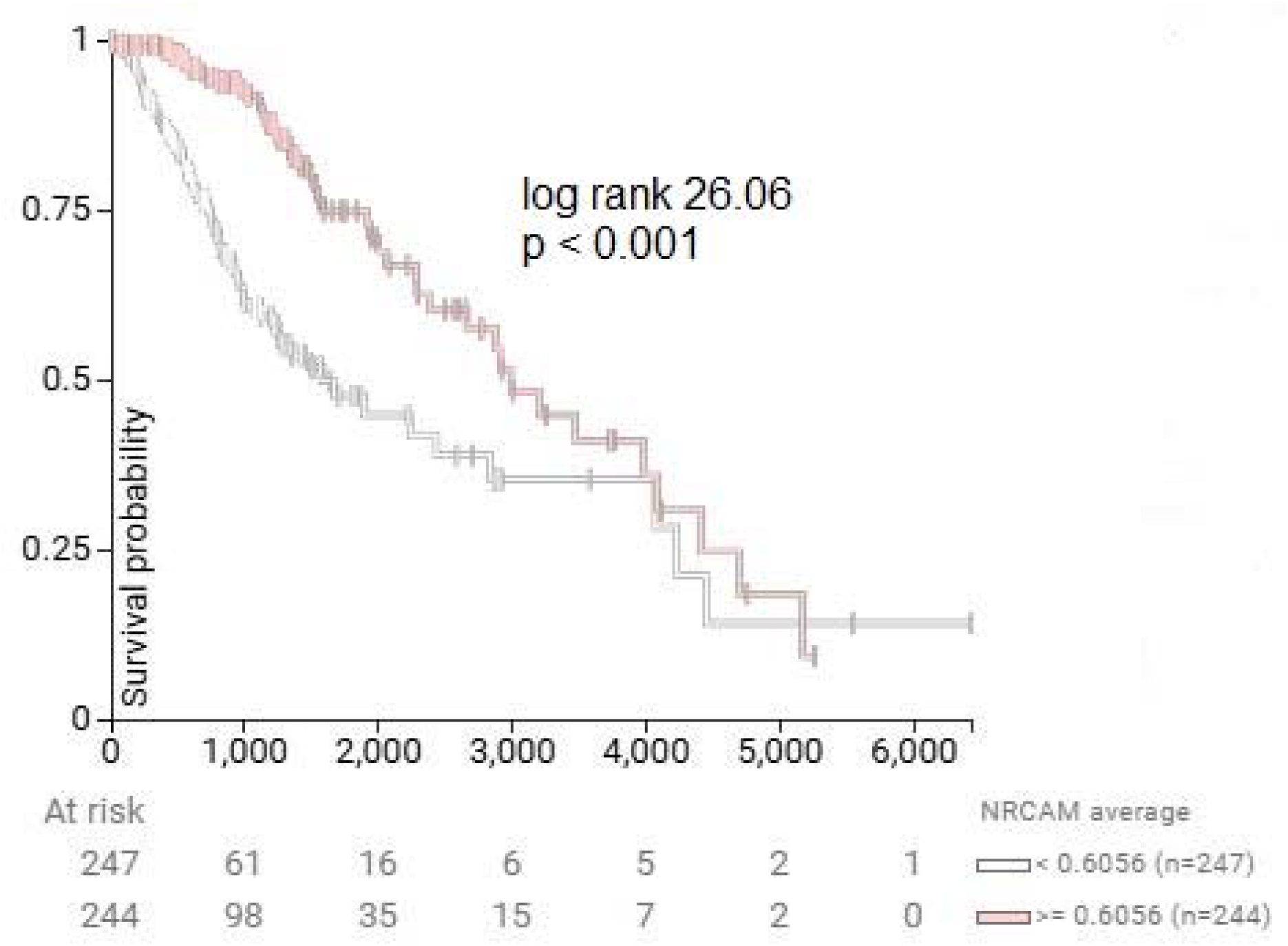
Survival probability versus NRCAM average methylation. More methylated tumors (upper curve) had significantly better survival.

No survival associations were observed for the small subset of patients with NRCAM mutations.

Integration of mutation count and genome-wide copy number burden further highlighted the rarity of NRCAM mutations. Scatterplots of mutation count versus fraction of the genome altered showed that NRCAM alterations did not correlate with global mutation burden (Figure 5).

**Figure 5.**
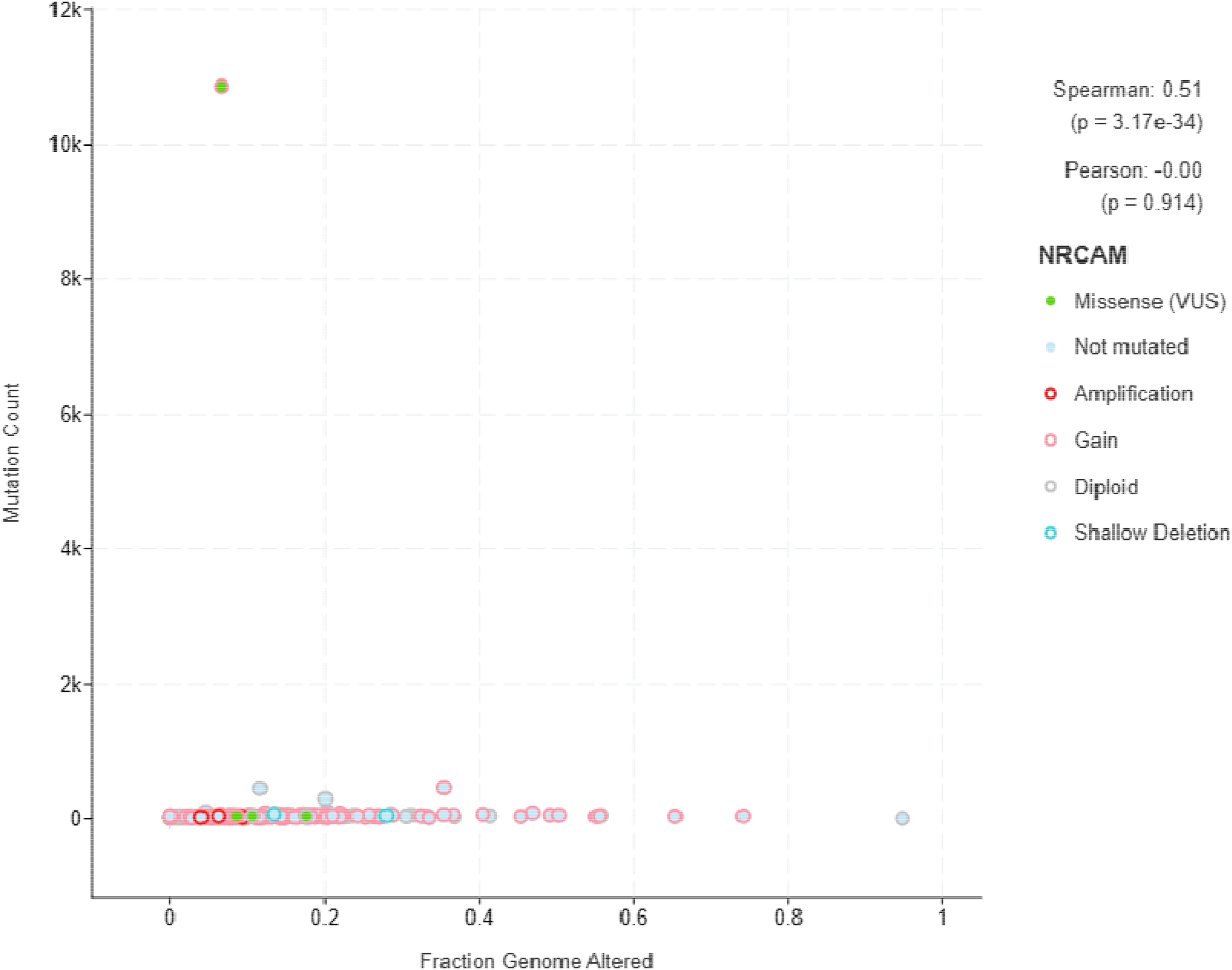
Scatterplots of NRCAM mutation count versus fraction of the genome altered. NRCAM alterations did not strongly correlate with FGA, suggesting that a sample with a high number of NRCAM mutations isn’t more likely to have a large portion of its genome altered, and vice versa. The Spearman correlation is likely inflated by a single outlier, reinforcing that there is no meaningful relationship between the two variables.

Overall, these results suggest that while mutations are rare, both copy number alterations and methylation changes at the NRCAM locus are detectable in gliomas and stratify patients by clinical outcome in TCGA cohorts.

## Discussion

In this study, we systematically examined NRCAM genetic and epigenetic alterations in gliomas using TCGA data. Our results demonstrate that while somatic mutations in NRCAM are rare, copy number variations and methylation changes are frequent and stratify patients by clinical outcome. Specifically, copy number gains were associated with shorter survival, whereas higher methylation of NRCAM correlated with improved prognosis. These findings suggest that epigenetic regulation and copy number imbalance, rather than coding mutations, may represent the most biologically relevant modes of NRCAM dysregulation in gliomas.

Our results complement and extend the observations reported by Sehgal et al., who identified alternative splicing events in NRCAM associated with glioma biology, especially involving amino acids encoded by exons 5 and 19, which are essential for high grade glioma growth [4]. Whereas Sehgal’s analysis implicated isoform-level diversity and splicing deregulation as potential drivers, our TCGA analysis highlights copy number and methylation as the predominant genomic alterations. Together, these findings suggest a broader model in which NRCAM dysregulation occurs through multiple layers of control—including splicing, copy number imbalance, and methylation shifts—that may converge on altered adhesion, signaling, and neural differentiation pathways in gliomas.

NRCAM is a neural cell adhesion molecule implicated in axon guidance, neuronal migration, and synaptic plasticity. Dysregulation of cell adhesion pathways has long been recognized as a hallmark of tumor invasion. The association of NRCAM copy number gains with poorer survival may reflect enhanced invasive potential and adaptability of glioma cells. Conversely, hypermethylation of NRCAM, associated with better survival, could represent transcriptional silencing of oncogenic isoforms or reduced cell adhesion signaling. These hypotheses align with Sehgal et al.’s demonstration of splicing-driven isoform differences, reinforcing the idea that NRCAM is a multi-layered regulator of glioma progression.

The clinical relevance of these findings lies in the potential utility of NRCAM alterations as biomarkers for glioma prognosis. NRCAM methylation status could serve as a non-invasive epigenetic biomarker measurable from circulating tumor DNA, providing prognostic stratification complementing existing classifiers such as IDH mutation and MGMT promoter methylation. Moreover, the convergence of copy number, methylation, and splicing dysregulation underscores NRCAM as a potential therapeutic target. Interventions aimed at modulating NRCAM signaling or isoform usage—whether through small molecules, epigenetic therapies, or splice-switching oligonucleotides— could represent innovative strategies for glioma management. Prospective validation in independent cohorts and functional studies will be required to move NRCAM from a correlative marker to a clinically actionable target.

Several limitations of this study must be acknowledged. First, TCGA datasets, while comprehensive, are retrospective and heterogeneous in terms of sample collection and clinical annotation. This limits the precision of correlative analyses between molecular alterations and outcomes. The retrospective nature of TCGA datasets and the correlative nature of our findings underscore the need for prospective validation in independent patient cohorts and functional studies to establish causality. Second, methylation measurements were probe-based and may not capture regulatory regions outside the Illumina array design. Third, while copy number and methylation alterations stratify survival, these associations are correlative; functional validation in cell and animal models will be necessary to establish causality. Finally, the integration of splicing data with TCGA copy number and methylation profiles was not feasible within the current analysis but represents an important direction for future studies, particularly considering Sehgal et al.’s findings.

In conclusion, this study provides evidence that NRCAM is recurrently dysregulated in gliomas through copy number alterations and methylation changes, which correlate with patient outcomes. When considered alongside the splicing alterations reported by Sehgal et al., our findings suggest that NRCAM dysregulation operates at multiple regulatory levels in gliomas. Future work should focus on integrating splicing, epigenetic, and genomic data in unified models, and on experimentally validating NRCAM’s functional role in glioma progression. This might be done, for example, by looking for associations between NRCAM methylation status and the expression of RNA-binding proteins like PTBP1 and SRRM4 that were discussed in the Sehgal et al. paper. This would provide a concrete, testable hypothesis for future research. Such studies may uncover novel therapeutic avenues targeting adhesion and signaling pathways in aggressive gliomas.

## Data Availability

All data produced are available online at https://xenabrowser.net/heatmap/
https://www.cbioportal.org/

https://xenabrowser.net/heatmap/

## Notes

### Competing Interest Statement

The authors have declared no competing interest.

### Funding Statement

This study did not receive any funding

### Author Declarations

The study used (or will use) ONLY openly available human data that were originally located at:https://xenabrowser.net/heatmap/ https://www.cbioportal.org/

